# PERIPHERAL BLOOD AS TOOL TO DETERMINE GENE EXPRESSION PATTERNS IN PATIENTS WITH PSYCHIATRIC, NEUROLOGICAL AND OTHER COMMON DISORDERS: A SYSTEMATIC REVIEW AND META-ANALYSIS PROTOCOL

**DOI:** 10.1101/19007633

**Authors:** Alana Castro Panzenhagen, Alexsander Alves-Teixeira, Martina Schroeder Wissmann, Carolina Saibro Girardi, Lucas Santos, Alexandre Kleber Silveira, Daniel Pens Gelain, José Cláudio Fonseca Moreira

## Abstract

**Introduction:** Common diseases are influenced by a variety of factors that can enhance one person’s susceptibility to developing a specific condition. Complex traits have been investigated in several biological levels. One that reflects the high interconnectivity and interaction of genes, proteins and transcription factors is the transcriptome. In this study, we disclose the protocol for a systematic review and meta-analysis aiming at summarizing the available evidence regarding transcriptomic gene expression levels of peripheral blood samples comparing subjects with psychiatric, neurological and other common disorders to healthy controls.

**Methods and analysis:** The investigation of the transcriptomic levels in the peripheral blood enables the unique opportunity to unravel the etiology of common diseases in patients *ex-vivo*. However, the experimental results should be minimally consistent across studies for them to be considered as the best approximation of the true effect. In order to test this, we will systematically identify all transcriptome studies that compared subjects with common disorders to their respective control samples. We will apply meta-analyses to assess the overall differentially expressed genes throughout the studies of each condition.

**Ethics and dissemination:** The data that will be used to conduct this study are available online and have already been published following their own ethical laws. Therefore this study requires no further ethical approval. The results of this study will be published in leading peer-reviewed journals of the area and also presented at relevant national and international conferences.

**Strengths and limitations of this study:**

➣ We present a new and systematically centered method to assess the overall effect of transcriptomic levels in the blood of subjects with common conditions.
➣ Meta-analyses are a robust statistical method to assess effect sizes across studies.
➣ The analysis is limited by the availability of studies, as well as their quality and comprehensiveness.
➣ Subgroup and meta-regression analyses will be also limited by the amount and quality of sample characterization variables made available by original studies.

## INTRODUCTION

Common human diseases are generally multifactorial, being influenced by both environmental and genetic factors that may interact on producing a certain phenotype [1,2]. Those diseases, related to the central nervous system (CNS) or otherwise, are usually associated with inflammation as well, which raises the possibility of systemic causal or resulting effects [3–9]. In neurodegenerative diseases as Parkinson’s or Alzheimer’s this association has even inspired the coining of the term “inflammaging” [10–12].

Diseases as diabetes, hypertension, psychiatric disorders or even continuous phenotypes, as body mass index (BMI) and height, are usually classified under the umbrella of complex traits. Those traits are influenced by an ensemble of different genetic variants and are therefore polygenic and “complex” [1,13]. This theory had already been partially developed by Fisher in 1919 [14]. Fisher’s “infinitesimal model” takes into account that the contribution of each variant and gene becomes smaller as the number of genes associated with a trait grows larger, resulting in a normally distributed phenotype [15].

More recently, researchers have proposed that complex traits are not only polygenic, but also “omnigenic”. The hypothesis of the omnigenic model states that all genes expressed in disease-relevant cells can actually affect core disease-related genes because of highly interconnected regulatory networks. This idea would suggest that a great amount of the heritability of those traits can also be explained by the influence of genes outside core pathways [13]. While this theory is nonetheless important to understand the etiology of complex traits, when referring ourselves to common diseases this should be translated into something that can be also applied therapeutically. In this sense, the continuing research on which pathways are mainly responsible for common diseases susceptibility remains crucial.

Multi-omics data has been vastly produced in the last few years from different types of human tissue and in several biological levels, such as in the genome, transcriptome, proteome, and even the microbiome. All these layers interact with each other and assessing a full perspective of the omics data has been one of the greatest challenges of today’s bioinformatics and biostatistics research [16–18]. However, the evidence is not even consolidated or provenly consistent across different studies within each omic. One of the most robust and widely applied statistical methods to solve similar problems in epidemiology is meta-analyzing data subsequently to a well-conducted systematic review of the literature in order to estimate overall effect sizes [19]. Having in mind that the interacting protein networks that influence diseases are mainly related to the regulation of gene expression, we have chosen to focus our study on the transcriptome level. Moreover, as tissue sample availability that is representative of common diseases is scarce, due to high heterogeneity and complexity, the investigation of peripheral factors becomes imperative. This is especially true for brain disorders and, as stated before, systemic effects might bring valuable insight to the matter. Hence, we have chosen to investigate transcriptomic differences in the most abundantly collected peripheral tissue: blood; which will hopefully solve inconsistencies across the field.

This study, therefore, aims at summarizing the available evidence regarding transcriptomic gene expression levels of blood samples comparing subjects with psychiatric, neurological and other common disorders to healthy controls. With this approach, we intend to identify the overall scenario regarding mRNA expression levels profile of complex traits, hopefully clarifying major inconsistencies in the field. Here we disclose the protocols for a systematic review and meta-analysis with the purpose of standardizing our methodology in order to increase quality and comprehensiveness, making it possible to be meticulously replicated hereafter.

## METHODS AND ANALYSIS

For the present study report, we have followed the Preferred Reporting Items for Systematic Reviews and Meta-Analyses Protocols (PRISMA-P) [20].

### Eligibility criteria

We will include studies that compared the gene expression levels in blood of patients with psychiatric, neurological, and other common disorders with healthy controls. We will include experimental studies that assessed gene expression levels by coding mRNA quantification through both microarray and RNA-sequencing methodology. Only studies with a specified control or resilience group will be considered. We will include randomized clinical trials (RCTs), cohort, and / or case-control studies, provided that they present a control group without specific and unique treatment administration. Studies should have collected blood samples (leukocytes, lymphocytes, peripheral blood mononuclear cells (PBMCs)) from patients affected with the disorder and healthy (or resilient) controls. All types of diagnosis will be included regardless of diagnosis manual or tool. Nevertheless, the diagnosis manual/tool will be included as variables in later sensitivity analyses. We have not imposed any restrictions regarding publication date, language or methodological quality, as well as age, sex or ethnicity of participants.

The following exclusion criteria will be applied: i) studies with a sample that comprises only genetically related individuals (*e*.*g*. family-based studies); ii) studies without original data; iii) studies in which every case has a unique type of comorbidity; iv) studies without healthy (or resilient) controls; v) studies in which remitters are referred to as healthy controls; vi) studies in which the blood samples received any kind of treatment *ex vivo* before microarray analysis or RNA-sequencing; and vii) studies in which all the case sample was receiving a specific type of drug that is not listed as “treatment as usual”, such as the mainly prescripted drug or class of drugs.

### Search and study identification

Studies will be identified through an online search using two different electronic databases: Gene Expression Omnibus (GEO) (https://www.ncbi.nlm.nih.gov/gds) and ArrayExpress (https://www.ebi.ac.uk/arrayexpress/). No search filters will be used and the reference list of published studies that were included will be searched for additional independent records. A further database search will be conducted for studies published since the last search date in GEO.

Search strategies will include subject headings for each disorder/disease (**Table 1**), indication for studies with human subjects, and blood. Full search strategies will be provided upon the final publication of the completed study.

**Table 1.** Common conditions and phenotypes that will be included in the study.

| **Psychiatric disorders** | **Neurological disorders** | **Immune-related diseases** | **Common diseases and phenotypes** |
| --- | --- | --- | --- |
| Attention-deficit/hyperactivity disorder | Alzheimer's disease* | Allergy | Body mass index* |
| Anorexia | Epilepsy | Asthma | Coronary artery disease* |
| Anxiety disorders | Stroke | Celiac disease | Diabetes* |
| Autism Spectrum Disorder* | Migraine disorders | Crohn's disease | Height* |
| Bipolar disorder | Multiple sclerosis | Lupus | Intelligence quotient |
| Major depressive disorder* | Parkinson's disease* | Psoriasis | Myocardial infarction |
| Obsessive compulsive disorder |  | Rheumatoid arthritis | Schooling years |
| Post-traumatic stress disorder |  |  |  |
| Schizophrenia* |  |  |  |
| Tourette syndrome |  |  |  |

\*Phenotypes that will be used in the pilot study

### Study selection

Study selection and inclusion will be performed in two steps. Firstly through title screening evaluation and secondly, by full GEO / ArrayExpress description details perusal. Both will be conducted by at least two independent reviewers (ACP, ATT, MSW, CSG, LS or AKT). Disagreements will be resolved in consultation with a third reviewer (ACP or JCFM).

### Data collection

The descriptive data of each study will be extracted by at least two independent reviewers (ACP, ATT, MSW, CSG, LS or AKT). Discrepancies will be resolved by a third reviewer (ACP or JCFM). Data will be extracted as follows: description of studies (sample size, age mean and standard deviation, gender frequencies in the sample, ethnicity and country/region of origin, date of data collection, type of platform used for transcriptome analyses, diagnostic manual / tool, and data on medication or psychotherapy being used). The transcriptomic outcome data (raw and/or normalized gene expression data) will be assessed through the *GEOquery* R package [21] or by downloading directly from the GEO and/or ArrayExpress websites. Whenever there is missing data, the authors of the original studies will be contacted. If no response is received in two months, the study will be excluded from the analyses.

### Outcome measures

Our primary outcome measure is mRNA levels assessed through microarray (or any other kind of specific array) or by RNA-sequencing methods. Those measures are available in full in the online repositories, and we will, therefore, be able to have access to raw data most of the time. Normalized data will not be a problem in the analyses, as we can convert raw data into normalized if the information on which method was used is provided. Whenever only the mean and standard deviation/error values are provided, the meta-analysis will still remain possible after appropriate measurement conversions.

### Bias assessment

Risk of bias will be assessed by two independent raters (ACP and MSW) through the NIH Quality Assessment of Case-Control Studies tool [22]. Any discrepancies in the assessment will be resolved through discussion and, where necessary, by a third rater (JCFM). Studies are rated as presenting high, unclear or low risk of bias. The NIH Quality Assessment tool comprises 12 questions about methodology quality of original case-control studies. However, we chose not to evaluate items 9 to 11, since they are not an issue in transcriptomic studies. This is because the exposure assessment, in this case, is performed by the automatic measurement of microarray or RNA-sequencing of samples. Furthermore, publication bias will be assessed using funnel plots and Egger’s regression test [23].

### Data synthesis

Gene expression data will be collapsed into one variable for each disorder through the *MetaMA* R package [24] and the overall differentially expressed genes computed. Subgroup analyses will be conducted for every study descriptive characteristics (e.g. gender, age, diagnostic tool, etc.) in which groups of at least three studies are formed.

#### Heterogeneity and subgroup analyses

Heterogeneity between studies will be assessed using both the *χ*^2^ and the I^2^ tests, in which a p-value ≤0.1 will be considered statistically significant in the first, and I^2^ values of 25%, 50% and 75% will be considered as low, moderate and high heterogeneity, respectively. We will conduct separate analyses of studies in groups by gender, age, and ethnicity of subjects when the data is available. We will also separate studies by disorder/disease subtypes or symptom grouping when pertinent, and by diagnostic tool when the data is available. Whether studies present high heterogeneity, we will also perform meta-regression analyses when enough data is available.

#### Sensitivity analyses

Sensitivity analyses will be also performed in order to evaluate result differences related to the effects of individual or specific groups of studies. The following sensitivity analysis are planed: i) the jackknife method, a common procedure used to test the stability of the outcome after excluding one result at a time [25], ii) excluding studies without formal diagnostic criteria from the analyses, and iii) excluding studies presenting a concerning risk of bias (the 25% worst ranked studies). The exploratory objectives of our sensitivity analyses will be to i) observe if any study skews the overall result, ii) examining the impact of studies without formal diagnostic criteria, and iii) evaluate the impact of studies with a high risk of bias.

### Pilot study

Each step of the analyses described above will be primarily tested in a pilot study, with a selected set of conditions, which are indicated by an asterisk in **Table 1**. We have chosen this set ambitioning to have more data on a broad variety of common conditions. This pilot study was thought of with the purpose of detecting possible misdesigns in the methodology and spot eventual issues that may emerge during the execution of the systematic review and/or meta-analysis.

## Data Availability

The datasets generated during and/or analyzed during the current study are available on request from the corresponding author.

## ETHICS AND DISSEMINATION

The data that will be used to conduct this study are available online and have already been published following their own ethical laws. Therefore this study requires no further ethical approval. The results of this study will be published in leading peer-reviewed journals of the area and also presented at relevant national and international conferences.

## Authors’ contributions

ACP and JCFM conceived the study. ACP designed and drafted the protocol and ATT, MSW, CSG, LS, AKT, DPG, and JCFM critically revised it. ACP, ATT, MSW, CSG, LS, and AKT will conduct the paired study search and paired inclusion of studies. Data extraction will be performed by ACP, AAT, and MSW. ACP will perform the analyses and draft the systematic review and meta-analysis manuscript and all authors will revise it. All authors have contributed to and approved the final protocol paper, as well as agreed on submission to BMJ Open.

